# A domain-specific approach to characterizing falls efficacy post-stroke

**DOI:** 10.1101/2025.08.18.25333910

**Authors:** Grace K. Kellaher, Ryan T. Pohlig, Darcy S. Reisman, Jeremy R. Crenshaw

## Abstract

**Objective:** The purpose of this study was to demonstrate that the construct of falls efficacy, as measured by the Activities-Specific Balance Confidence (ABC) scale, is comprised of distinct factors in those with chronic stroke.

**Design:** This study is a cross-sectional analysis of pre-existing data from a research registry.

**Setting:** Academic institution.

**Participants:** 248 community-dwelling adults history of one or more strokes, verified by CT/MRI report or past provider.

**Interventions:** Not applicable.

**Main Outcome Measure(s):** Goodness-of-Fit indices (Root Mean Square Error of Approximation (RMSEA), Comparative Fit Index (CFI), and Tucker-Lewis Fit Index (TLI)).

**Results:** To test our hypothesis that the ABC scale is comprised of distinct factors linked to the balance domains, we proposed that the ABC scale items are indicative of three subdomains, 1) anticipatory control, 2) walking balance, and 3) reactive balance. A confirmatory factor analysis was performed to test this hypothesized structure. We assigned ABC Q1,2,8,10-12 to walking balance, Q3-7,9 to anticipatory control, and Q13-16 to reactive balance. All indicators significantly loaded as hypothesized. Modification indices suggested that Q6, which asks about standing on a chair and reaching for something, might be multidimensional and load onto both the anticipatory control and reactive balance latent constructs. Therefore, confidence in reaching while on a stool reflects confidence in both anticipatory and reactive balance.

**Conclusion(s):** These results suggest that the items in the ABC scale measure three subfactors that align with anticipatory control, walking balance, and reactive balance.

## Introduction

Falls efficacy has been defined as a person’s confidence in their ability to perform activities of daily living without falling.^1^ Individuals with chronic stroke have low falls efficacy,^2–4^ and up to 80% of these individuals with chronic stroke express a fear of falling.^5^ Falls efficacy is commonly quantified for older adults and neurologic populations in both clinical and laboratory settings using self-reported questionnaires such as the Activities-specific Balance Confidence (ABC) scale.^6^ For people with chronic stroke, the range of reported mean ABC scale scores^2–4,7–10^ (43.3-78.5 out of 100) extends below thresholds associated with high fall risk^11^ (ABC < 68) and activity avoidance^12^ (ABC < 80). Accordingly, addressing falls efficacy is an important component of post-stroke rehabilitation.

Physical-activity-based interventions only have a moderate beneficial effect on falls efficacy that is not sustained with time.^13^ A challenge with addressing falls efficacy after a stroke is that it is likely comprised of multiple balance domains. Physical therapy guidelines^14^ and the organization of clinical balance assessments^15^ define individual domains of balance as sensory orientation, anticipatory control, walking balance, and reactive balance. Similarly, the ABC scale has questions specific to the domains of anticipatory control, walking balance, and reactive balance. Although the questions of the ABC scale assess efficacy across distinct domains of balance, the questionnaire itself results in a single, composite measure of falls efficacy. A previous study of the ABC scale in people with chronic stroke supports the idea that measures of falls efficacy do not represent a single construct and is likely also comprised of multiple domains.^3^ This study concluded that the ABC scale consists of two factors, tasks of “low” and “high” perceived risk.

Factors categorized as high- and low-risk, however, have limited clinical utility, as they do not inform the specific balance interventions to improve falls efficacy. A domain-specific approach to measuring falls efficacy can specifically inform the balance skills to be targeted with intervention. This specificity is key to enabling skill mastery.^16^ Prior work has shown that the biomechanics and performance of balance tasks across the identified balance domains are not strongly correlated in those with chronic stroke.^17–20^ We know that skill mastery is one of the most effective ways to improve falls efficacy post-stroke.^21,22^ In a clinical trial of perturbation-based, reactive-balance training, benefits were only observed when focusing on the specific tests of reactive balance, and not on an aggregate score of tests across balance domains within the Mini-BESTest.^23^ The correlations between aggregate ABC scores and domain-specific balance function are not strong,^3,24–26^ and aggregate ABC scores only have moderate improvements with domain-specific balance interventions.^13^ Thus, a domain-specific approach to measuring falls efficacy will likely be more sensitive to domain-specific impairments and changes in balance.

Because falls efficacy measures are not specific to balance domains, they are limited in their capacity to inform intervention targets and detect benefits from domain-specific interventions. For that reason, the purpose of this study was to demonstrate that the construct of falls efficacy in those with chronic stroke is comprised of multiple factors that align with distinct domains of balance within the existing ABC scale. We hypothesized that the ABC scale would be comprised of factors linked to the domains of anticipatory control, walking balance, and reactive balance.

## Methods

### Participants

This study was a cross-sectional analysis of pre-existing data from 248 community-dwelling adults with stroke who are enrolled in a Stroke Research Registry at the university (Table 1). The Stroke Research Registry is an IRB-approved recruitment tool that tracks individuals with stroke who are interested in participating in research studies. To be included in the registry, participants must have been an adult with a history of one or more strokes, verified by CT/MRI report or past provider. All participants signed informed consent approved by the institution’s Human Subjects Institutional Review Board prior to participation.

**Table 1.**
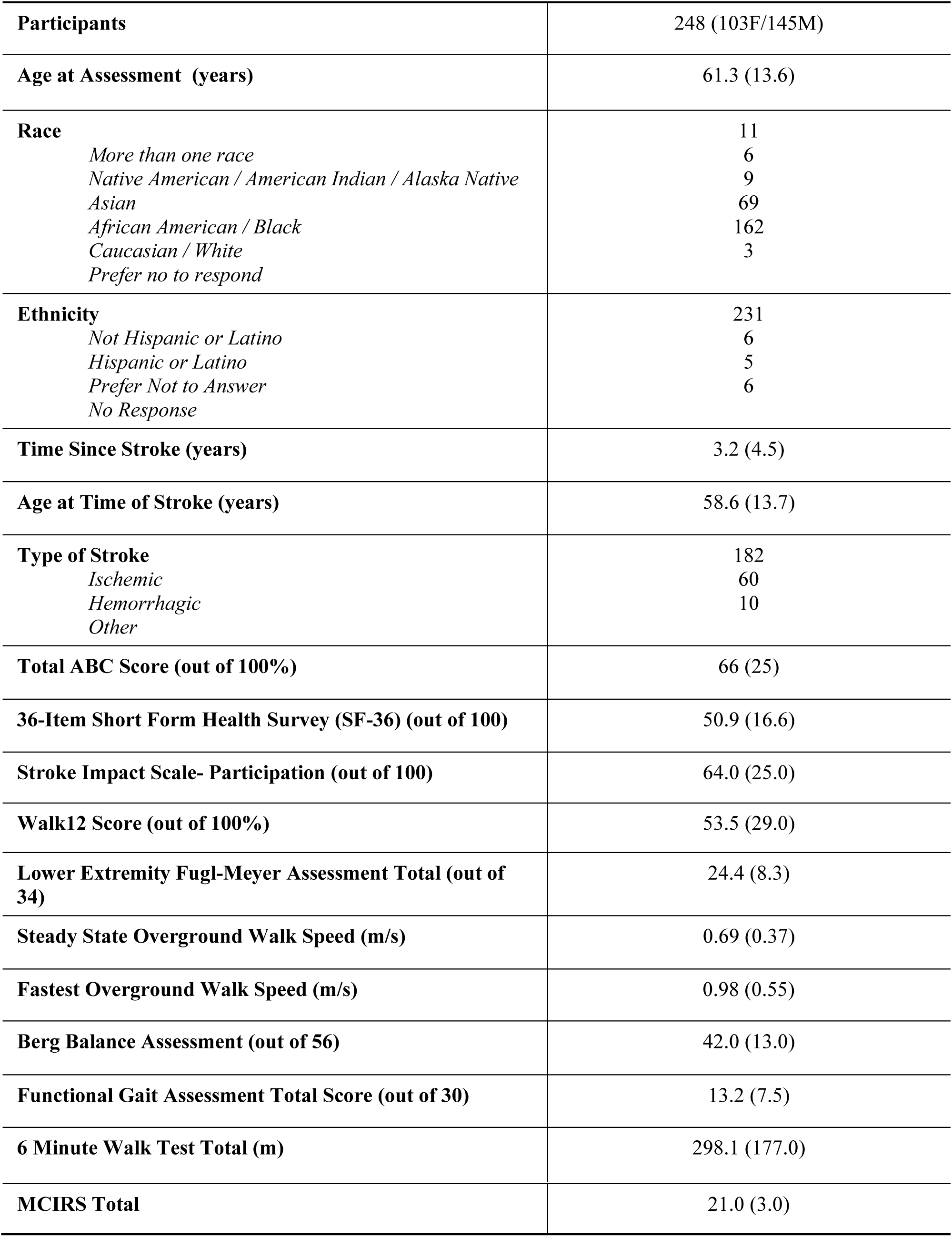
Participant demographics and physical function for 248 community-dwelling adults with stroke enrolled in the Stroke Studies Registry. Values are presented as mean (standard deviation).

|  |  |
| --- | --- |
| **Participants** | 248 (103F/145M) |
| **Age at Assessment (years)** | 61.3 (13.6) |
| **Race**<br>*More than one race*<br>*Native American / American Indian / Alaska Native*<br>*Asian*<br>*African American / Black*<br>*Caucasian / White*<br>*Prefer no to respond* | 11<br>6<br>9<br>69<br>162<br>3 |
| **Ethnicity**<br>*Not Hispanic or Latino*<br>*Hispanic or Latino*<br>*Prefer Not to Answer*<br>*No Response* | 231<br>6<br>5<br>6 |
| **Time Since Stroke (years)** | 3.2 (4.5) |
| **Age at Time of Stroke (years)** | 58.6 (13.7) |
| **Type of Stroke**<br>*Ischemic*<br>*Hemorrhagic*<br>*Other* | 182<br>60<br>10 |
| **Total ABC Score (out of 100%)** | 66 (25) |
| **36-Item Short Form Health Survey (SF-36) (out of 100)** | 50.9 (16.6) |
| **Stroke Impact Scale- Participation (out of 100)** | 64.0 (25.0) |
| **Walk12 Score (out of 100%)** | 53.5 (29.0) |
| **Lower Extremity Fugl-Meyer Assessment Total (out of 34)** | 24.4 (8.3) |
| **Steady State Overground Walk Speed (m/s)** | 0.69 (0.37) |
| **Fastest Overground Walk Speed (m/s)** | 0.98 (0.55) |
| **Berg Balance Assessment (out of 56)** | 42.0 (13.0) |
| **Functional Gait Assessment Total Score (out of 30)** | 13.2 (7.5) |
| **6 Minute Walk Test Total (m)** | 298.1 (177.0) |
| MCIRS Total | 21.0 (3.0) |

### Protocol

Upon enrollment in the registry, participants completed a one-time, standardized assessment conducted by a licensed physical therapist. The assessment included recording demographic information, characteristics about their stroke, and a battery of clinical tests and surveys. These surveys included the ABC scale,^6^ which is comprised of 16 questions that are each answered with a numerical value of 0-100, with a score of 100 indicating complete confidence in a person’s balance ability and a score of 0 indicating no confidence in a person’s balance ability. When administered to those with stroke, the questionnaire has acceptable internal consistency (Cronbach’s α = 0.94) and test-retest reliability (ICC = 0.85).^3^

### Data Management

Study data were collected and managed using REDCap electronic data capture tools hosted at our institution.^27,28^ REDCap (Research Electronic Data Capture) is a secure, web-based software platform designed to support data capture for research studies, providing 1) an intuitive interface for validated data capture; 2) audit trails for tracking data manipulation and export procedures; 3) automated export procedures for seamless data downloads to common statistical packages; and 4) procedures for data integration and interoperability with external sources.

### Framework

To test our hypothesis that the ABC scale will be comprised of distinct factors linked to the balance domains, we proposed that the ABC scale items can be categorized, as follows (Table 1): *Anticipatory Control:* Tasks that necessitate motor adjustments before and during volitional changes in posture.^14^

*Walking Balance:* Tasks that are characterized primarily by walking with no external, mechanical perturbations, such as walking around the house.

*Reactive Balance:* Tasks that require a rapid sensorimotor response to prevent a loss of balance.^14^

These tasks all involve an external perturbation, like a bump or moving support surface, or the implied acceleration of the base of support, such as walking on ice.

The 16 items on the ABC scale were assigned to a balance domain by the study team. We acknowledge that ABC scale items may have features across multiple balance domains. The deciding factors when hypothesizing a categorization of these items were to 1) align them in similar domains as the activities in the Mini-BESTest,^15^ and 2) use our expert judgment to predict which domain of exercise intervention would have the most benefit on that item’s performance. For example, perturbation-based balance training^29–31^ should most benefit the efficacy of tasks categorized as reactive balance.

### Statistical Analysis

We conducted a confirmatory factor analysis (CFA) using the ABC, hypothesizing that the 16 items come from three subdomains, “anticipatory control,” “walking balance,” and “reactive balance.” The hypothesized model’s indicators and their corresponding latent constructs are shown in Table 2. Evaluation of the overall model fit was carried out using the chi-squared goodness of fit, the Root Mean Square Error of Approximation (RMSEA), Comparative Fit Index (CFI), and Tucker-Lewis Fit Index (TLI). Standardized parameter estimates are reported and hypotheses about specific pathways were evaluated using z-tests. All analyses were performed with α = 0.05.

**Table 2.** Activities-specific Balance Confidence (ABC) Scale Items.

| *“How confident are you that you will <u>not</u> lose your balance or become unsteady when you...”* |  |
| --- | --- |
| Context | Proposed Balance Domain |
| *Q1...walk around the house?* | walking |
| *Q2...walk up or down stairs?* | walking |
| *Q8...walk outside the house to a car parked in the driveway?* | walking |
| *Q10...walk across a parking lot to the mall?* | walking |
| *Q11...walk up or down a ramp?* | walking |
| *Q12...walk in a crowded mall where people rapidly walk past you?* | walking |
| *Q3...bend over and pick up a slipper from the front of a closet floor?* | anticipatory |
| *Q4...reach for a small can off a shelf at eye level?* | anticipatory |
| *Q9...get into or out of a car?* | anticipatory |
| *Q7...sweep the floor?* | anticipatory |
| *Q5...stand on your tip toes and reach for something above your head?* | anticipatory |
| *Q6...stand on a chair and reach for something?* | anticipatory |
| *Q13...are bumped into by people as you walk through the mall?* | reactive |
| *Q14...step onto or off of an escalator while holding onto a railing?* | reactive |
| *Q15...step onto or off an escalator while holding onto parcels such that you cannot hold onto the railing?* | reactive |
| *Q16...walk outside on icy sidewalks?* | reactive |

A sample size calculation was performed for this study using Preacher & Coffman’s method.^32^ The total number of parameters to be estimated for the model is 35 (16 factor loadings, 16 disturbances, 3 correlations among factors), leaving 101 degrees of freedom. For n = 188, power is greater than 0.95 with df = 101, for testing model misfit comparing RMSEA of 0.11 (moderate fit), compared to 0.05 (close fit), for α = 0.05.

## Results

The mean ABC total score for our sample was 66% (SD = 25%). The mean score for ABC items related to walking balance (Q1,2,8,10,11,12) was 76% (SD = 31%). The mean score for ABC items related to anticipatory control (Q3,4,5,6,7,9) was 69% (SD = 37)%. The mean score for ABC items related to reactive balance (Q13, 14,15,16) was 48% (SD = 39%). Mean scores for individual questions can be seen in Figure 1.

**Figure 1.**
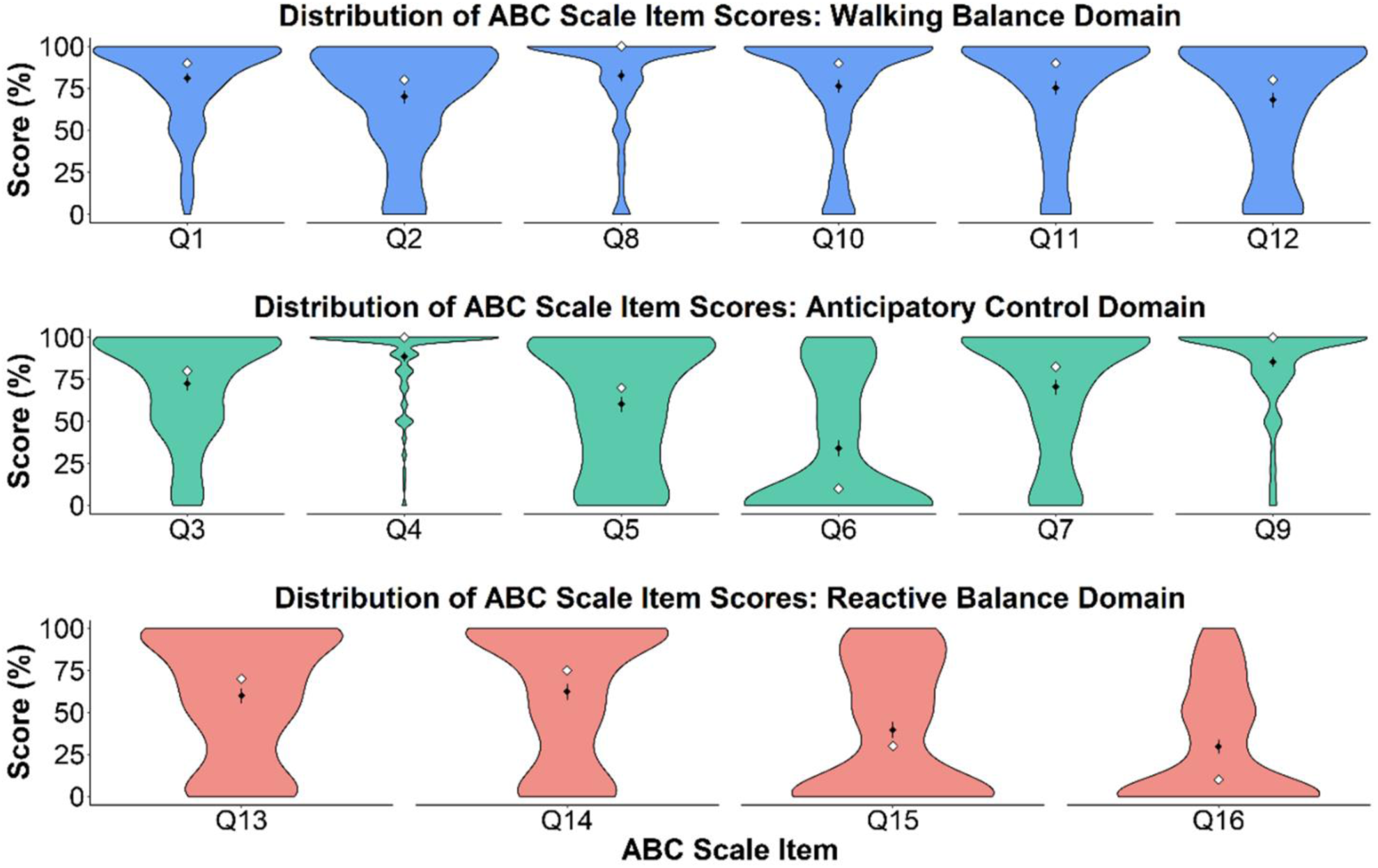
Distribution of individual ABC question scores for 248 individuals with chronic stroke. Black diamonds represent mean scores for each question, error bars represent the 95% CI, white diamonds represent median scores for each question.

In the confirmatory factor, all indicators significantly loaded as hypothesized (p<0.05). Questions 1, 2, 8, 10, 11, and 12 loaded onto the latent construct of “walking balance.” Questions 3, 4, 5, 6, 7, and 9 loaded onto the latent construct of “anticipatory control.” Questions 13, 14, 15, and 16 loaded onto the latent construct of “reactive balance.” The model showed adequate fit (RMSEA = 0.118, CFI = 0.846, TLI = 0.818, SRMR = 0.070) (Figure 2).

**Figure 2.**
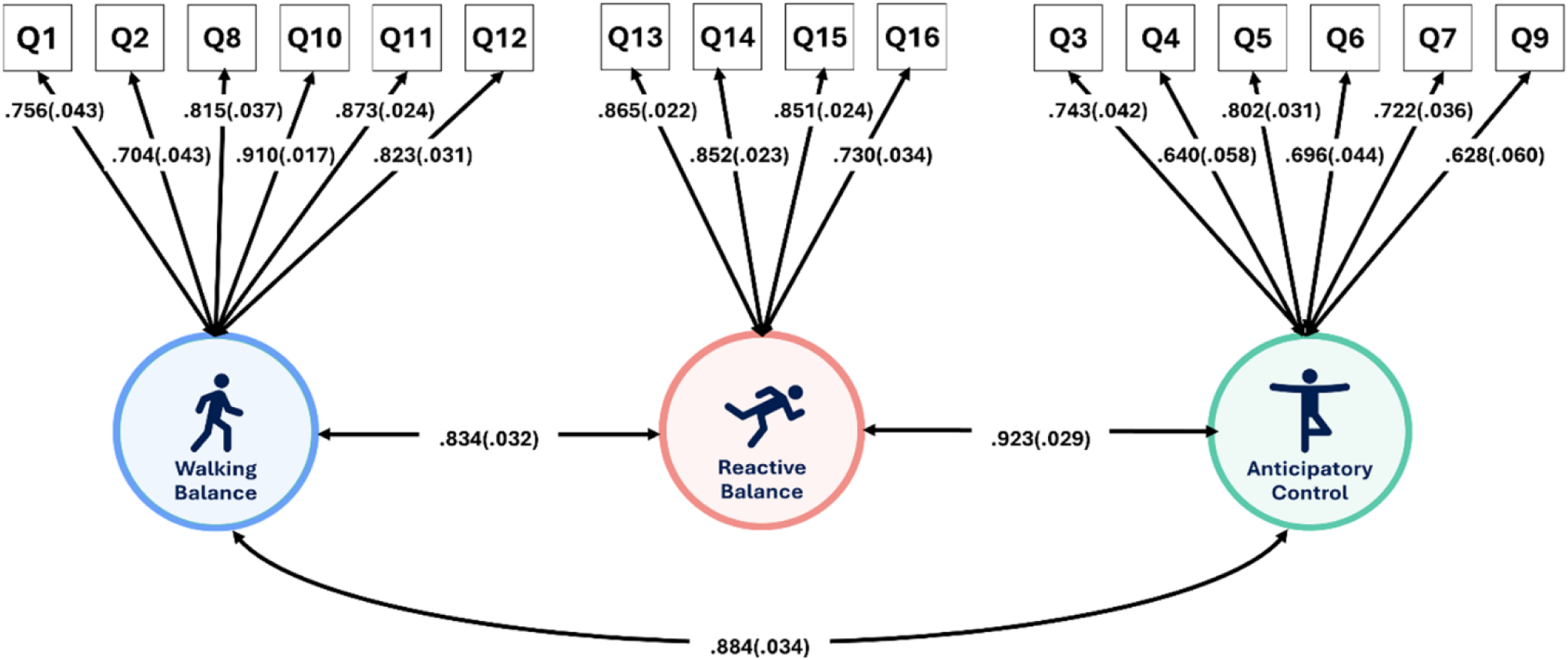
Results of initial CFA. All indicators significantly loaded as hypothesized; Q1,2,8,10-12 on walking, Q3-7,9 on anticipatory control, and Q13-16 on reactive balance (p<0.05). The model showed adequate fit (RMSEA=0.118, CFI=0.846, TLI=0.818, SRMR=0.070).

Modification indices suggested that Question 6, “How confident are you that you will not lose your balance or become unsteady when you…stand on a chair and reach for something” might be multidimensional. Our team initially hypothesized this item to be associated with “anticipatory control” because the primary action of this task is reaching for an object. However, participants may reply with their efficacy in regaining balance should they become unsteady while standing on the chair. Such unsteadiness may occur frequently in those with stroke, given their altered planning and execution of a standing reach.^33^ Such unsteadiness would require the skill of “reactive balance”, and the elevated height of the chair would increase the cost of a failed reactive response. Given this potential role of reactive balance, a subsequent CFA was performed with Question 6 to load onto both the “anticipatory control” and “reactive balance” latent constructs. This resulted in an improved model fit from our initial CFA (RMSEA = 0.109, CFI = 0.869, TLI = 0.843, SRMR = 0.064) (Figure 3).

**Figure 3.**
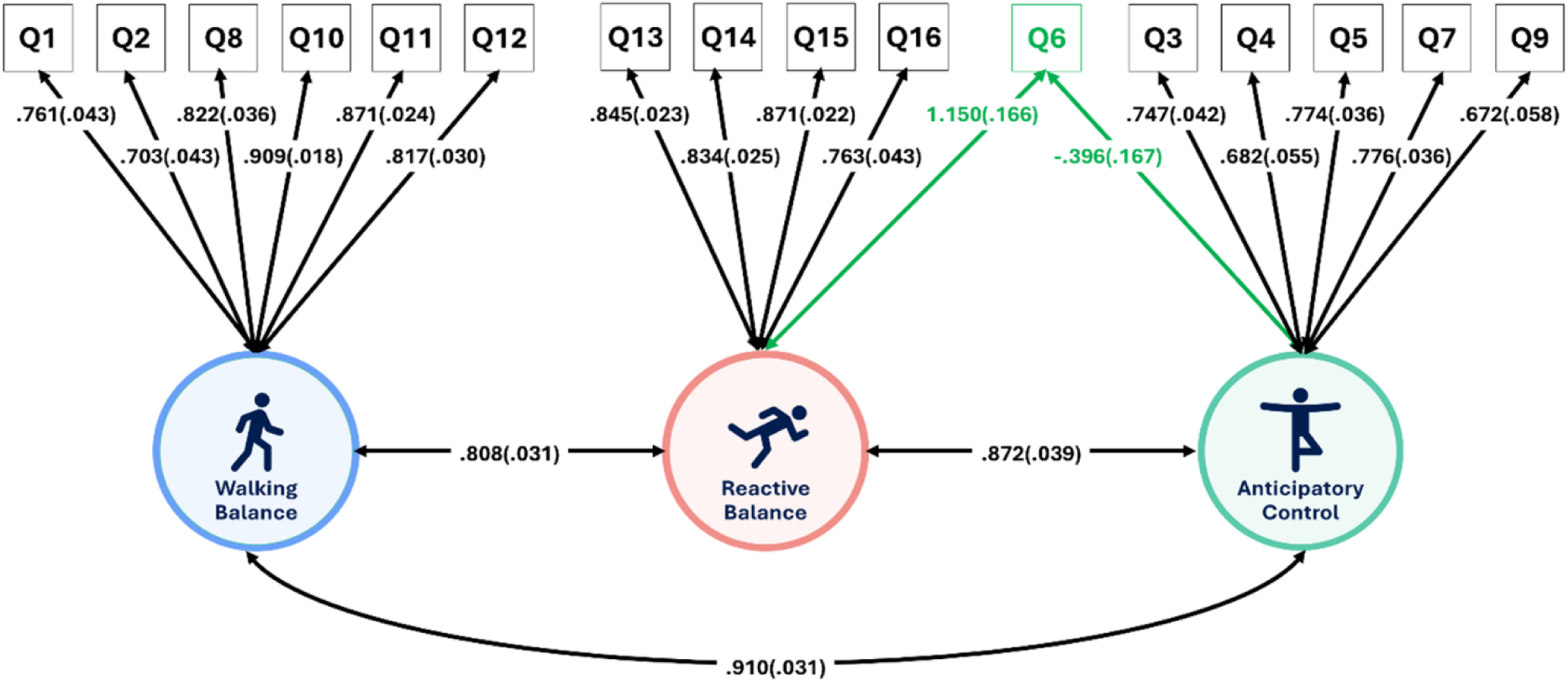
Results of post-hoc CFA, allowing Q6 to load onto “anticipatory control” and “reactive balance.” This analysis resulted in an improved model CFA (RMSEA=0.109, CFI=0.869, TLI=0.843, SRMR=0.064).

Lastly, to validate our model we conducted an additional CFA on ABC questionnaire data from a separate set (n = 119, Table 3) of individuals enrolled in the Stroke Studies Registry, who had participated in a multi-site clinical trial aimed at improving real-world walking activity after a stroke (NIH R01HD086362), collected pre-intervention. In this model, we again hypothesized that Questions 1, 2, 8, 10, 11, and 12 would load onto the latent construct of “walking balance.”

**Table 3.** Participant demographics and physical function of 119 community-dwelling adults with stroke enrolled in the Stroke Studies Registry who had participated in a multi-site clinical trial aimed at improving real-world walking activity after a stroke. Values are presented as mean (standard deviation).

|  |  |
| --- | --- |
| **Participants** | 248 (103F/145M) |
| **Age at Assessment (years)** | 61.3 (13.6) |
| **Race**<br>*More than one race*<br>*Native American / American Indian / Alaska Native*<br>*Asian*<br>*African American / Black*<br>*Caucasian / White*<br>*Prefer no to respond* | 11<br>6<br>9<br>69<br>162<br>3 |
| **Ethnicity**<br>*Not Hispanic or Latino*<br>*Hispanic or Latino*<br>*Prefer Not to Answer*<br>*No Response* | 231<br>6<br>5<br>6 |
| **Time Since Stroke (years)** | 3.2 (4.5) |
| **Age at Time of Stroke (years)** | 58.6 (13.7) |
| **Type of Stroke**<br>*Ischemic*<br>*Hemorrhagic*<br>*Other* | 182<br>60<br>10 |
| **Total ABC Score (out of 100%)** | 66 (25) |
| **36-Item Short Form Health Survey (SF-36) (out of 100)** | 50.9 (16.6) |
| **Stroke Impact Scale- Participation (out of 100)** | 64.0 (25.0) |
| **Walk12 Score (out of 100%)** | 53.5 (29.0) |
| **Lower Extremity Fugl-Meyer Assessment Total (out of 34)** | 24.4 (8.3) |
| **Steady State Overground Walk Speed (m/s)** | 0.69 (0.37) |
| **Fastest Overground Walk Speed (m/s)** | 0.98 (0.55) |
| **Berg Balance Assessment (out of 56)** | 42.0 (13.0) |
| **Functional Gait Assessment Total Score (out of 30)** | 13.2 (7.5) |
| **6 Minute Walk Test Total (m)** | 298.1 (177.0) |
| **MCIRS Total** | 21.0 (3.0) |

Questions 3, 4, 5, 6, 7, and 9 would load onto the latent construct of “anticipatory control.” Questions 13, 14, 15, 16, and 6 would load onto the latent construct of “reactive balance.” This model had a similar fit to our original model (RMSEA = 0.103, CFI = 0.847, TLI = 0.816, SRMR = 0.073a and all indicators significantly loaded as hypothesized (p<0.05) except for Question 6, which was unidemsional and only loaded on “anticipatory control.” These results suggest that our model is reproducible.

## Discussion

The purpose of this study was to demonstrate that the construct of falls efficacy, as measured by the ABC scale, is comprised of distinct factors in individuals with chronic stroke. We hypothesized that the ABC scale would be comprised of distinct factors linked to the balance domains of anticipatory control, walking balance, and reactive balance. In support of our hypothesis, the results of our CFA demonstrated that our proposed measurement model of distinct factors exhibited good fit to our data, supporting our hypothesized relationship between the items on the ABC scale and the factors of anticipatory control, walking balance, and reactive balance (Figure 2). Each of the items on the ABC scale loaded highly onto its respective latent factor. In our post-hoc analysis, allowing for cross-loading of ABC Question 6 onto both the latent factors of anticipatory control and reactive balance improved the model fit, suggesting that Question 6 on the ABC scale is multidimensional and represents both anticipatory control and reactive balance constructs of falls efficacy (Figure 3).

Our findings that the ABC scale is comprised of multiple factors in individuals with chronic stroke aligns with previous work on using the ABC scale in individuals with chronic stroke that found falls efficacy is not a single construct.^3^ Our findings, however, indicate that the different constructs represented in the ABC scale align with the domains of balance. Balance domain-specific scores of falls efficacy may have more clinical utility than a composite score or a “high” versus “low” risk categorization established in previous work.^3^ Similar to the way domain-specific diagnoses of balance dysfunction aid clinicians in determining the most appropriate interventions to improve balance ability,^14^ domain-specific scores for falls efficacy may be more beneficial for detecting the balance domains where individuals post-stroke have the lowest falls efficacy. Given that falls efficacy can be improved through skill mastery,^16,34^ measures of falls efficacy that are balance domain-specific may allow clinicians to prescribe more appropriate, skill-specific balance interventions for individuals post-stroke.

Falls efficacy is a determinant of activity, engagement, and participation in the free-living environment after a stroke.^4,9,35–41^ Prior work has shown that aggregate measures of falls efficacy mediate the relationship between walking capacity (i.e., walking speed and endurance) and walking activity, as well as the relationship between the environment and walking activity, in those who have experienced a stroke.^38,39,42^ Thus, low falls efficacy may be one reason the benefits of improved walking capacity after rehabilitation intervention do not translate to more walking activity in the free-living environment. We do not yet know, however, the specific domains of falls efficacy that influence walking activity, nor do we know if interventions that improve such domain-specific measures subsequently benefit walking activity. Future work should investigate the relationship between the domains of falls efficacy and walking activity, as we may be able to target specific features of walking activity through a domain-specific approach to improving balance and its corresponding self-efficacy.

### Limitations

There are some limitations to using the ABC scale as our measure of falls efficacy in people with chronic stroke. The ABC scale instructions specify not “losing balance” or “becoming unsteady”. This wording is ambiguous in the context of reactive balance. Tasks that involve reactive balance, such as being bumped into, are inherently characterized by a loss of balance or unsteadiness. We assume that participants reply with their efficacy in regaining balance or steadiness after the perturbation (i.e. reactive balance). This ambiguous wording of the ABC scale items related to reactive balance may limit the validity of that domain.

The ABC scale is also limited in how it characterizes the domains of falls efficacy, and the domains of falls efficacy represented in the ABC scale may represent task difficulty, rather than balance skill. For example, the walking tasks described in the ABC scale items do not examine biomechanically challenging circumstances, which may limit the scale’s sensitivity to detecting impairments in walking balance efficacy. Other measures of falls efficacy, such as the Modified Gait Efficacy Scale or the Falls Efficacy Scale-International may provide more challenging circumstances of walking. Additionally, recent work has expanded the definition of falls efficacy to include a person’s “perceived ability to manage a threat of a fall”.^43^ The ABC scale does not account for efficacy related to safe landing or getting off of the ground after a fall occurs. These aspects may be better characterized by other scales that include items related to balance recovery self-efficacy, such as the Falls Efficacy Scale-International, which asks about “concern” for falling, or the Balance Recovery Scale, which measures perceived reactive balance recovery, although it has not yet been validated in individuals with chronic stroke.

Lastly, we did not create a factor aligning with the “sensory orientation” aspect included in the Mini-BESTest^23^. Those aspects are assessed with standing postures under different visual or proprioceptive constraints. No questions on the ABC are similar to these static tasks. It is likely that sensory orientation is an aspect that underlies the efficacy of the more dynamic conditions of walking, anticipatory control, and reactive postural control. Therefore, it may not need to be characterized as a separate construct of efficacy.

## Conclusion

Overall, we have confirmed that the ABC scale consists of three latent factors that align with the specific balance domains of anticipatory control, walking balance, and reactive balance. This distinction can allow us to develop domain-specific measures of falls efficacy that may be more sensitive to detecting the benefits of domain-specific balance interventions.

## Acknolwedgments

We thank Michael Christensen for his assistance with the data management, organization, and submission to the NICHD DASH for this study.

## Acknowledgment of Prior Presentation

This work was previously presented in a poster format at the 2025 American Society for Neurorehabilitation Annual Meeting on April 24, 2025 in Atlanta, Georgia, USA.

## Acknowledgement of Financial Support

Research reported in this publication was supported by the Eunice Kennedy Shriver National Institute of Child Health & Human Development of the National Institutes of Health under Award Number R03HD113864. The content is solely the responsibility of the authors and does not necessarily represent the official views of the National Institutes of Health.

## Conflict of Interest

None

## Abbreviation Definition

ABC: Activities-specific Balance Confidence
Q: question

## Data Availability

All data produced in the present study are available upon reasonable request to the authors. All data produced will become available online through the NICHD Data and Specimen Hub (DASH) once approved.

## Notes

### Competing Interest Statement

The authors have declared no competing interest.

### Author Declarations

The Human Subjects Institutional Review Board of the University of Delaware gave ethical approval for this work.

